# Accuracy of deep learning based computed tomography diagnostic system of COVID-19: a consecutive sampling external validation cohort study

**DOI:** 10.1101/2020.11.15.20231621

**Authors:** Tatsuyoshi Ikenoue, Yuki Kataoka, Yoshinori Matsuoka, Junichi Matsumoto, Junji Kumasawa, Kentaro Tochitatni, Hiraku Funakoshi, Tomohiro Hosoda, Aiko Kugimiya, Michinori Shirano, Fumiko Hamabe, Sachiyo Iwata, Shingo Fukuma, Japan COVID-19 AI team

## Abstract

**Objectives:** Ali-M3, an artificial intelligence, analyses chest computed tomography (CT) and detects the likelihood of coronavirus disease (COVID-19) in the range of 0 to 1. It demonstrates excellent performance for the detection of COVID-19 patients with a sensitivity and specificity of 98.5 and 99.2%, respectively. However, Ali-M3 has not been externally validated. Our purpose is to evaluate the external validity of Ali-M3 using Japanese sequential sampling data.

**Methods:** In this retrospective cohort study, COVID-19 infection probabilities were calculated using Ali-M3 in 617 symptomatic patients who underwent reverse transcription-polymerase chain reaction (RT-PCR) tests and chest CT for COVID-19 diagnosis at 11 Japanese tertiary care facilities, between January 1 and April 15, 2020.

**Results:** Of 617 patients, 289 patients (46.8%) were RT-PCR-positive. The area under the curve (AUC) of Ali-M3 for predicting a COVID-19 diagnosis was 0.797 (95% confidence intervals [CI]: 0.762-0.833) and goodness-of-fit was P = 0.156. With a cut-off of probability of COVID-19 by Ali-M3 diagnosis set at 0.5, the sensitivity and specificity were 80.6% and 68.3%, respectively, while a cut-off of 0.2 yielded a sensitivity and specificity of 89.2% and 43.2%, respectively. Among 223 patients who required oxygen support, the AUC was 0.825 and sensitivity at a cut-off of 0.5 and 0.2 were 88.7% and 97.9%, respectively. Although the sensitivity was lower when the days from symptom onset were few, sensitivity increased for both cut-off values after 5 days.

**Conclusions:** Ali-M3 was evaluated by external validation and shown to be useful to exclude a diagnosis of COVID-19.

**Key Points:**

1. The area under the curve (AUC) of Ali-M3, which is an AI system for diagnosis of COVID-19 based on chest CT images, was 0.797 and goodness-of-fit was P = 0.156.
2. With a cut-off of probability of COVID-19 by Ali-M3 diagnosis set at 0.5, the sensitivity and specificity were 80.6% and 68.3%, respectively, while a cut-off of 0.2 yielded 89.2% and 43.2%.
3. Although low sensitivity was observed in less number of days from symptoms onset, after 5 days high increasing sensitivity was observed. In patients requiring oxygen support, the AUC was higher that is 0.825.

## Introduction

A proper triage system is necessary during this coronavirus disease (COVID-19) pandemic era,[1, 2] as improper triage systems may disadvantage patients and lead to wastage of personal protective equipment (PPE) and hospital infections through admission of infected patients to facilities, causing collapse of the medical system. Although reverse transcription-polymerase chain reaction (RT-PCR) tests have been developed, the delay in waiting for RT-PCR results can hamper proper triage.

Computed tomography (CT) is a fast and useful diagnostic tool. Some studies have reported the characteristic findings on chest CT images of COVID-19 patients.[3-8] Use of chest CT images by radiologists has shown high diagnostic performance for COVID-19. However, even radiologists’ interpretations vary largely, because of the influence of their habituation in the interpretation of COVID-19 CT images.[9] Therefore, using CT as a diagnostic tool in general clinical practice is difficult in the current situation.

Diagnostic support systems using artificial intelligence (AI) have the potential to replace many of the routine detection, characterisation, and quantification tasks currently performed by radiologists using cognitive ability.[10] AI can prevent the variability of diagnosis from inter- and intra-reader variability. In China, where COVID-19 infection originated, many AI systems were developed for establishing a diagnosis of COVID-19 based on chest CT images.[11-15] One such system, Ali-M3, can detect the likelihood of COVID-19 in the range of 0 to 1, and has excellent accuracy for the detection of COVID-19 with an accuracy, sensitivity, and specificity of 99.0, 98.5, and 99.2%, respectively. Although Ali-M3 has excellent accuracy, it was developed in a virtual population, which consisted of 3,067 examinations for COVID-19; 1,996 for community-acquired pneumonia; and 1,975 for non-pneumonia, which was different from the general population and its accuracy could be overestimated.[16]

To use Ali-M3 to diagnose exclusion of COVID-19, its external validity must be evaluated based on the distribution of diseases in a real-world setting. We here conducted a retrospective cohort study to evaluate the external validity of Ali-M3 using the Japanese sequential sampling data of patients who underwent RT-PCR tests and chest CT for diagnosis of COVID-19.

## Materials and Methods

### Study design

This retrospective cohort study consisted of 11 Japanese tertiary care facilities that provided treatment for COVID-19 in each area. We partially followed the guidelines of the Transparent Reporting of a Multivariable Prediction Model for Individual Prognosis or Diagnosis Statement to plan and report this study (Supplemental Table 1).[17] The institutional review board of each facility approved the study and the need to obtain written informed consent was waived.

**Table 1.** Demographics of patients’ characteristics.

| Variable | Symptomatic patients | Patients using oxygen support | Asymptomatic patients |
| --- | --- | --- | --- |
| N | 617 | (223) | (86) |
| Age (years old) <sup>+</sup> | 59.6 (19.2) | 68.3 (16.4) | 54.5 (22.4) |
| Sex (Male) | 377 (61.2) | 158 (70.9) | 40 (46.5) |
| Real-time PCR test (Positive) | 289 (46.8) | 97 (43.5) | 37 (43.0) |
| Body temperature ($\geq 37^{\circ}$) | 391 (66.5) | 143 (69.8) | |
| Systolic Blood Pressure ($\leq 90$ mmHg) | 18 (3.2) | 11 (5.2) | |
| Pulse ($\geq 120$ bpm) | 48 (8.2) | 22 (10.2) | |
| Respiratory rate ($\geq 25$ /minute) | 92 (20.5) | 64 (38.3) | |
| Saturation of percutaneous oxygen ($\leq 92$ %) | 105 (17.7) | 62 (28.7) | |
| Oxygen use | 223 (36.1) | 223 (100.0) |  |
| Vasopressor use | 14 (2.3) | 14 (6.3) |  |
| Distribution of symptoms reported |  |  |  |
| Dry cough | 232 (37.6) | 67 (30.0) |  |
| Chills | 91 (14.7) | 40 (17.9) |  |
| Sore throat | 159 (25.8) | 38 (17.0) |  |
| Diarrhea | 66 (10.7) | 17 (7.6) |  |
| Joint or muscle pain | 46 (7.5) | 12 (5.4) |  |
| Conjunctivitis | 30 (4.9) | 9 (4.0) |  |
| Loss of smell or taste | 55 (8.9) | 21 (9.4) |  |
| Exposure history |  |  |  |
| No | 484 (78.4) | 191 (85.7) | 62 (72.1) |
| Within family | 39 (6.3) | 11 (4.9) | 6 (7.0) |
| Other persons | 94 (15.2) | 21 (9.4) | 18 (20.9) |
| Any international travel | 44 (7.1) | 6 (2.7) | 9 (10.5) |
| Current Smoking | 99 (16.0) | 41 (18.4) | 11 (12.8) |
| Past medical history |  |  |  |
| Cardiac artery disease | 46 (7.5) | 24 (10.8) | 4 (4.7) |
| Stroke | 60 (9.7) | 34 (15.2) | 2 (2.3) |
| Chronic heart failure | 69 (11.2) | 43 (19.3) | 4 (4.7) |
| Chronic kidney disease | 58 (9.4) | 33 (14.8) | 7 (8.1) |
| Chronic obstructive pulmonary disease | 69 (11.2) | 34 (15.2) | 7 (8.1) |
| Malignancy | 105 (17.0) | 62 (27.8) | 8 (9.3) |
| Immune deficiency | 32 (5.2) | 17 (7.6) | 1 (1.2) |
| Hypertension | 119 (19.3) | 71 (31.8) | 11 (12.8) |
| Diabetes | 116 (18.8) | 64 (28.7) | 13 (15.1) |
| Any other disease | 188 (30.5) | 73 (32.7) | 29 (33.7) |

PCR, polymerase chain reaction; bpm, beats per minute
\*Patients using oxygen support were included in symptomatic patients
+ is continuous data and the others are count data. Continuous variables are expressed as mean (SD) and count data as number (percentage).

### Participants

We included patients who underwent both RT-PCR examinations and chest CT for the diagnosis of COVID-19. The potentially eligible participants were identified on the advice of physicians that both RT-PCR test and chest CT be obtained when the patients presented with symptoms or were suspected of having COVID-19. The detailed information of the inclusion criteria is shown in Supplemental Table 2. We selected patients by using consecutive sampling methods between January 1 and April 15, 2020. The RT-PCR results were extracted from the patients’ medical records at each facility. Patients were excluded when the time-interval between chest CT and the first RT-PCR assay was longer than 7 days.

**Table 2.**
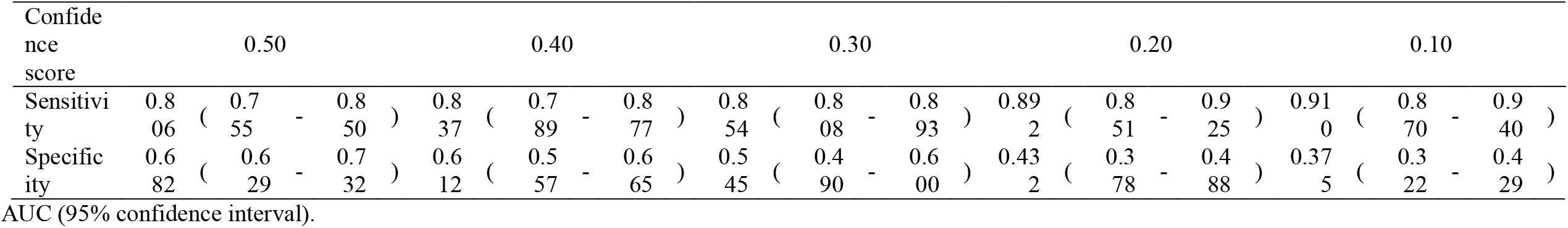
Moving cut-off confidence score and test performance.

All available data on the database were used to maximize the power and generalizability of the results.

### Chest CT protocols

All images were obtained on one of five types of CT systems, with the patient in the supine position.

The details of scanning parameters and systems are shown in Supplemental Table 3.

### Image analysis

We used a three-dimensional deep learning framework for the detection of COVID-19 infections.[16] The details of this model are included in Appendix 1. The learning of Ali-M3 was stopped before our evaluation. We set a cut-off point for the model output at 0.5, because this cut-off point was used during the developing stage. The investigators who entered the CT images data into Ali-M3 were blinded to the RT-PCR results.

### Reference standard

The diagnosis of COVID-19 was established by the RT-PCR test, which detected the nucleic acid of severe acute respiratory syndrome coronavirus 2 (SARS-CoV-2) in the sputum, throat swabs, and secretions of the lower respiratory tract samples.[18] We established the RT-PCR tests as the main reference standard. Although the findings of chest CT, interpreted by radiologists, were included as the reference standard in the derivation study, we did not include it as the reference standard in the present study.

### Statistical analysis

Statistical analysis was performed using R statistical software, version 3.6.3 (R Foundation for Statistical Computing). Data analysis was performed in a complete-case dataset. Continuous variables are presented as means (standard deviation) and categorical variables are presented as counts and percentages. Using the RT-PCR results as reference, the area under the curve (AUC), sensitivity, specificity, positive-predictive value, and negative-predictive value of the likelihood of COVID-19 as derived from the Ali-M3’s analysis of chest CT imaging were calculated. A 95% confidence interval (CI) was determined by the Wilson score method. The goodness-of-fit was calculated using the Le Cessie-Van Houwelingen normal test statistic for the unweighted sum of squared errors.

### Sensitivity analysis

#### 1. Moving cut-off point

The objective of this study was to determine whether this AI model could be used as a screening tool for COVID-19 in the real world. In a clinical situation, physicians require an accurate diagnosis of COVID-19; hence, they insist more on sensitivity than on specificity. For sensitivity analysis, we moved the cut-off point and observed sensitivities and specificities to minimize overlooking COVID-19 patients.

#### 2. Simulation of imperfect reference

In the main analysis, we assumed RT-PCR as the perfect reference (100% sensitivity and 100% specificity). However, in the real world, RT-PCR is not the perfect reference since the sensitivity of the RT-PCR test was estimated at 0-80%.[19] To evaluate the effect of this imperfect reference, we calculated the sensitivity, specificity, and AUC of Ali-M3 using the methods and R code described in the Supplemental Material when varying the sensitivity, but fixing the specificity of RT-PCR at 100%.[20]

#### 3. Effect of the number of days after symptom onset

The number of days that have passed since the onset of symptoms affects the performance of antibody and RT-PCR tests in COVID-19 patients.[19, 21] However, it was not clear if this could affect CT images in COVID-19 patients. Sensitivity and specificity were calculated for a group of patients whose symptom onset date was known, among those were those with 14 days or more, as well as those at every 2 days from 0 to 13 days after symptom onset.

#### 4. Effect of symptom severity

Imaging is not routinely indicated as a screening test for COVID-19 in asymptomatic individuals.[22] However, CT images are used in assessment of disease severity. We established the severity by evaluating whether oxygen therapy was required and if the patient was asymptomatic while undergoing CT.

#### 5. Effect of reconstruction slice

The thickness of the reconstruction slice can affect the diagnostic performance.[23] We separated the dataset for the main analysis by a 3-mm reconstruction slice thickness to account for the fissure in our data set between 3 mm and 4 mm and calculated the performance of the model in each dataset.

## Results

### Study population characteristics

Figure 1 shows the patient flow diagram. Data of 749 patients were evaluated. We assessed 617 symptomatic patients in this validation study. The characteristics of the study population for the main analysis datasets are shown in Table 1. Overall, 289 patients (46.8%) were diagnosed with COVID-19 using the RT-PCR test. Thirteen patients need more than two RT-PCR tests before being diagnosed with COVID-19. Major symptoms were dry cough (37.6%), fever (33.5%), and sore throat (25.8%).

**Figure 1.**
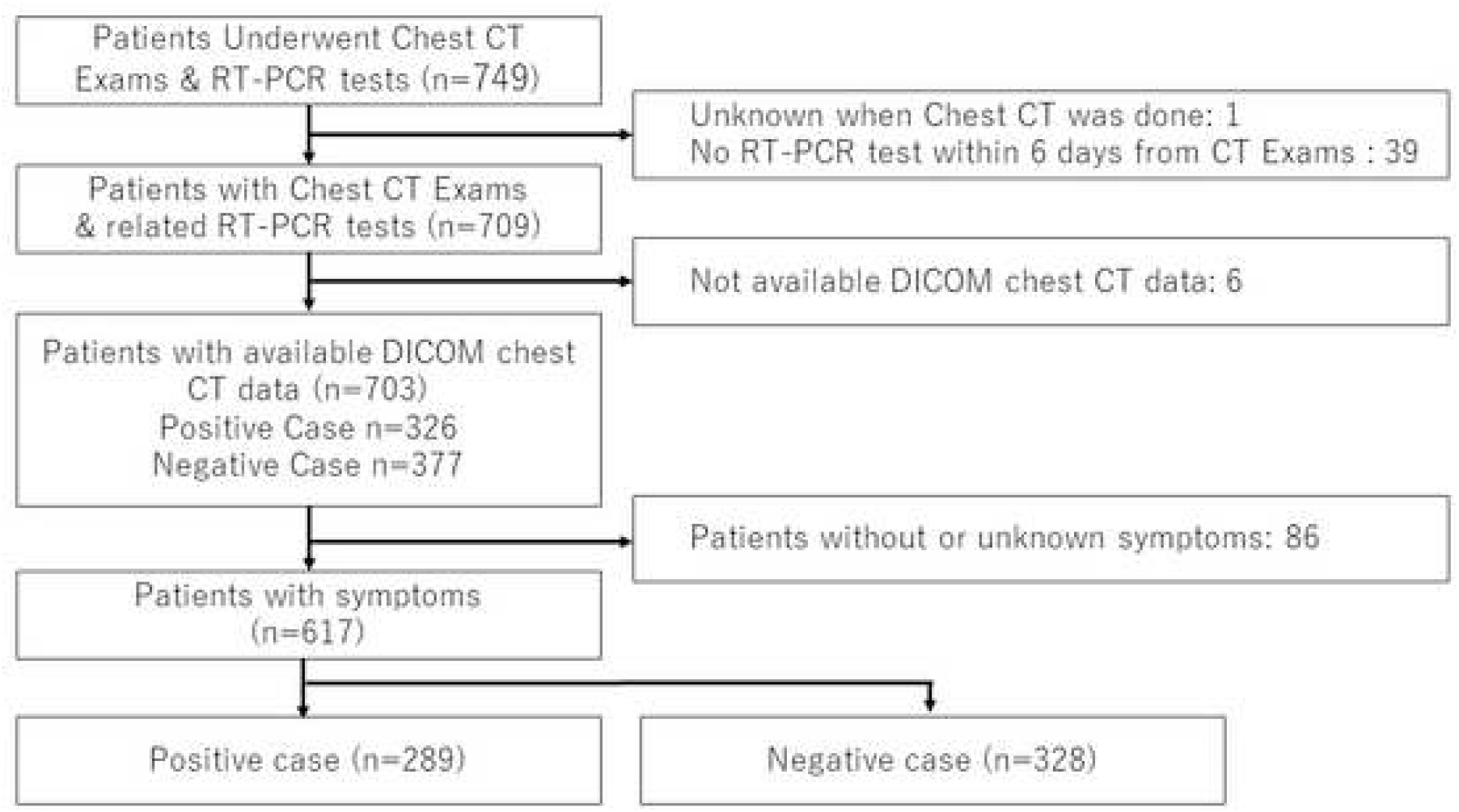
Patient flow. Abbreviations: CT, computed tomography; RT-PCR, reverse transcription polymerase chain reaction; DICOM, digital imaging and communications in medicine

### Model performance

The performance of the confidence score after validation among symptomatic patients is shown in Figure 2. The performance of the confidence score was *P* = 0.156 for the goodness-of-fit, and the AUC was 0.797 (95% CI 0.762-0.833). The relationship between the score and predicted probability is shown in Figure 2. The optimal cut-off point with maximal sensitivity and specificity was 0.5, and the sensitivity and specificity were 80.6% (233 of 289) [95% CI: 75.6-85.0%] and 68.3% (224 of 328) [95% sCI, 63.0-%], respectively.

**Figure 2.**
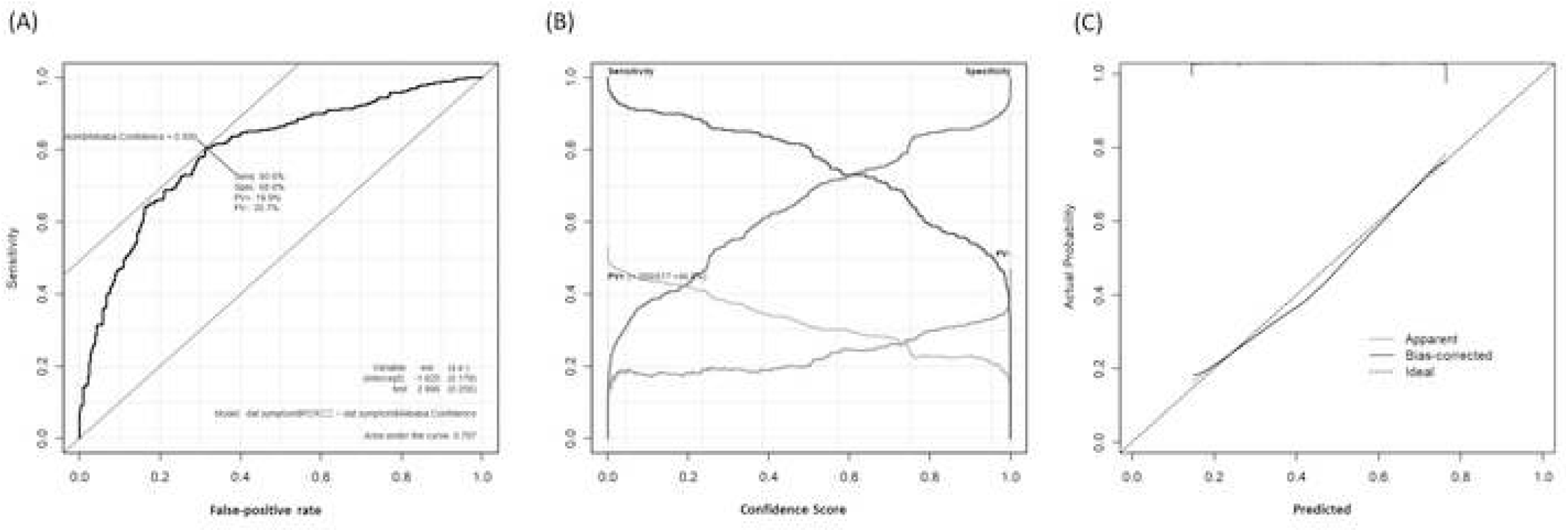
Differential performance of Ali-M3 for coronavirus disease in symptomatic patients. (A) A plot of test sensitivity (y-coordinate) versus its false-positive rate (x-coordinate) obtained at each cutoff level confidence score. The area under the receiver operating characteristic curve is 0.797 and the Youden index is 0.50. (B) A plot of test sensitivity, specificity, positive predictive value (PV+), and negative predictive value (PV-) in y-coordinate versus confidence score obtained from Ali-M3 in x-coordinate. The PV+ is dark gray and the PV-is light gray. The maximum PV+ is 46.8% and the maximum PV-is 53.2%. (C) This graph shows the goodness-of-fit. The dashed line is an ideal line that predicts the probability obtained from the confidence score of Ali-M3 equal to the actual probability. The pointed line is the fitted line that is estimated with non-linear assumption alone. The dashed line is the fitted line that is estimated with non-linear assumption and considering the bias in nonparametric estimation using the le Cessie-van Houwelingen method.

### Sensitivity analysis

#### 1. Moving cut-off point

Table 2 shows the relationship between cut-off points for the confidence score and performance.When the cut-off point was 0.2, the sensitivity and specificity were 89.2% and 43.3%, respectively.

#### 2. Simulation of imperfect reference

Figure 3 shows the sensitivity and specificity, with the assumption of imperfect reference of RT-PCR test. The AUC was 0.865. When the cut-off point was set at 0.5, using the Youden Index, the sensitivity and specificity were 80.6% and was 81.3%, respectively. When the cut-off point was set at 0.2, the sensitivity and specificity were 89.2% and 51.9%, respectively.

**Figure 3.**
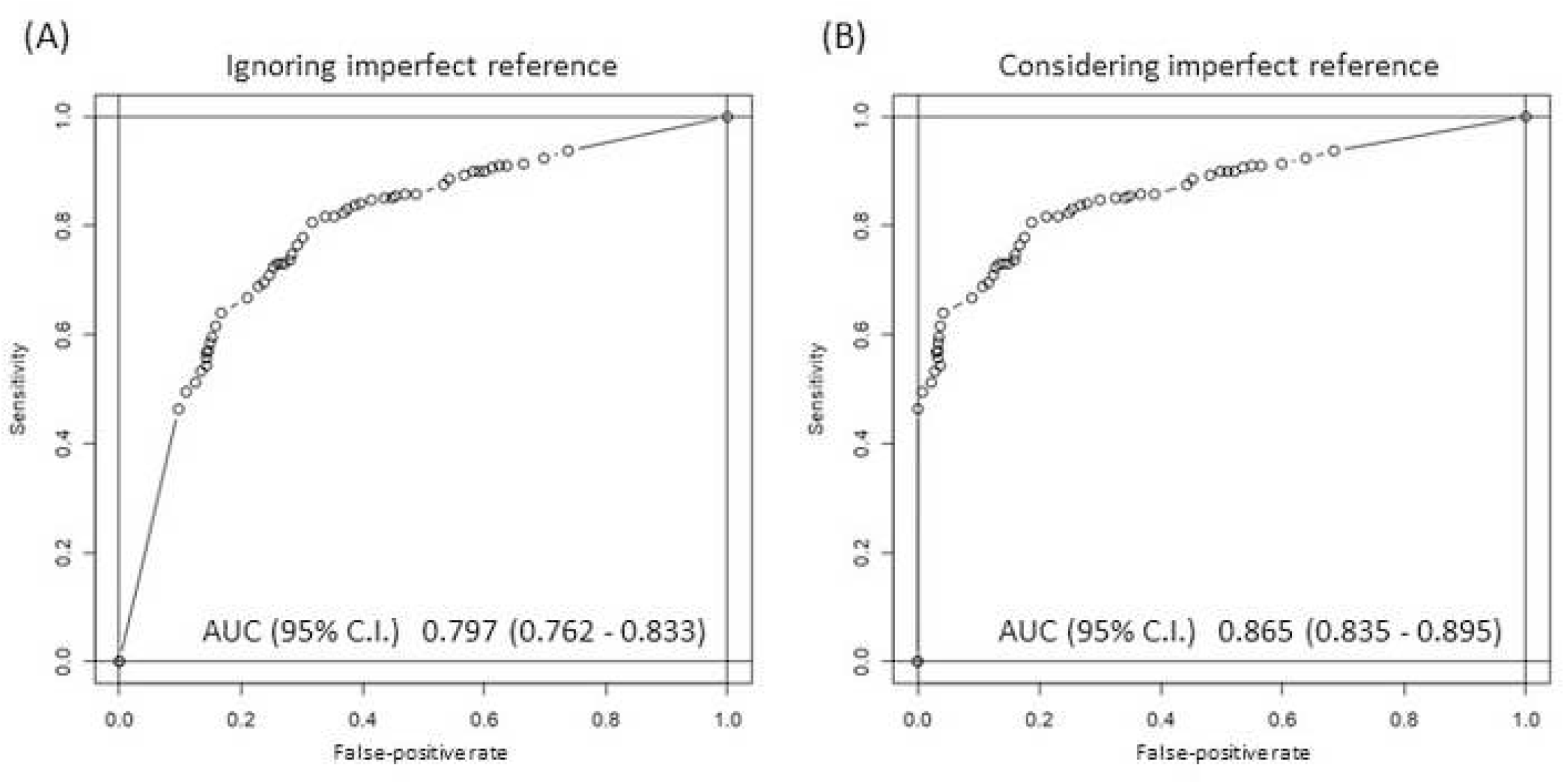
Relationship between test performance and the number of days after the onset of symptoms. (A) The graph shows the relationship between test performance and the number of days after the onset of symptoms when the confidence score from Ali-M3 is at 0.20. (B) The graph shows the relationship between test performance and the number of days after onset of symptoms when the confidence score from Ali-M3 is at 0.50. Light gray bar shows the number of patients included in the strata of days after the onset of symptoms, following the right axis. One stratum includes 2 days from day 0 to day 13. The stratum to the extreme right includes 14 days or more. Following the left axis, solid lines are sensitivity in strata and dash lines are specificity in strata.

#### 3. Effect of number of days after symptom onset

Of all symptomatic patients, 600 patients (97.2%) were included in this sensitivity analysis. Of these, the number of days after the onset of symptoms was not known for 17 patients. Figure 4 shows the relationship between test performance and the number of days since the onset of symptoms when the confidence score of Ali-M3 was set at 0.5 or 0.2. Sensitivity values started at 0.7 and increased up to 1.0 until 10-11 days in both cases. However, specificity values remained similar across the strata. The sensitivity increased over 0.9 when the confidence score was set at 0.2 than when the confidence score was set at 0.5.

**Figure 4.**
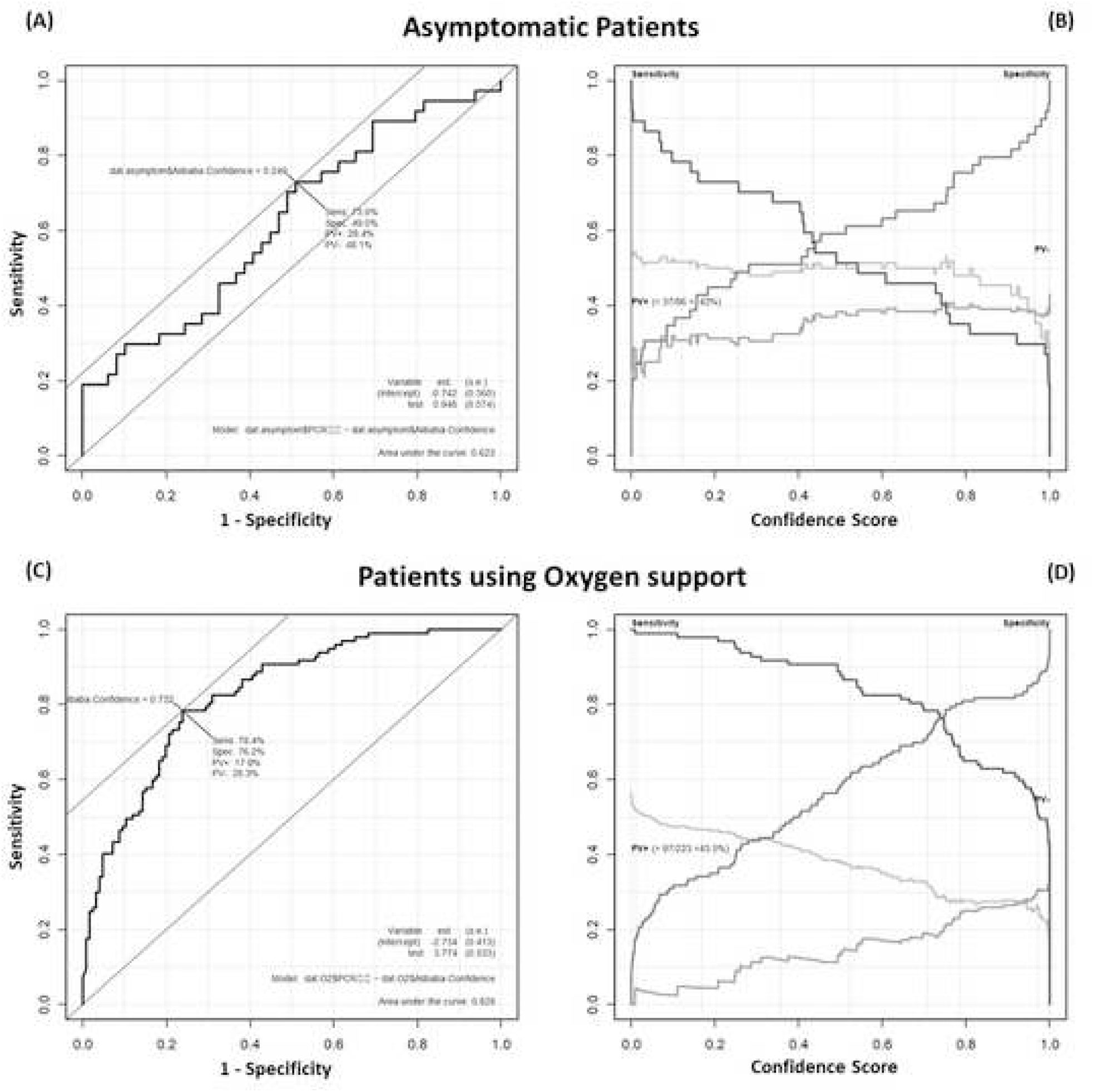
Receiver operating characteristic (ROC) curves in ignoring imperfect reference and considering imperfect reference. (A) A plot of test sensitivity (y-coordinate) versus its false-positive rate (x-coordinate) obtained at each cutoff level confidence score ignoring imperfect reference. The area under the ROC curve is 0.797. (B) A plot of test sensitivity (y-coordinate) versus its false-positive rate (x-coordinate) obtained at each cutoff level confidence score considering imperfect reference. The area under the ROC curve is 0.865.

#### 4. Changing the eligibility criteria

The effects of changing the criteria for patient eligibility are shown n Figure 5.

**Figure 5.**
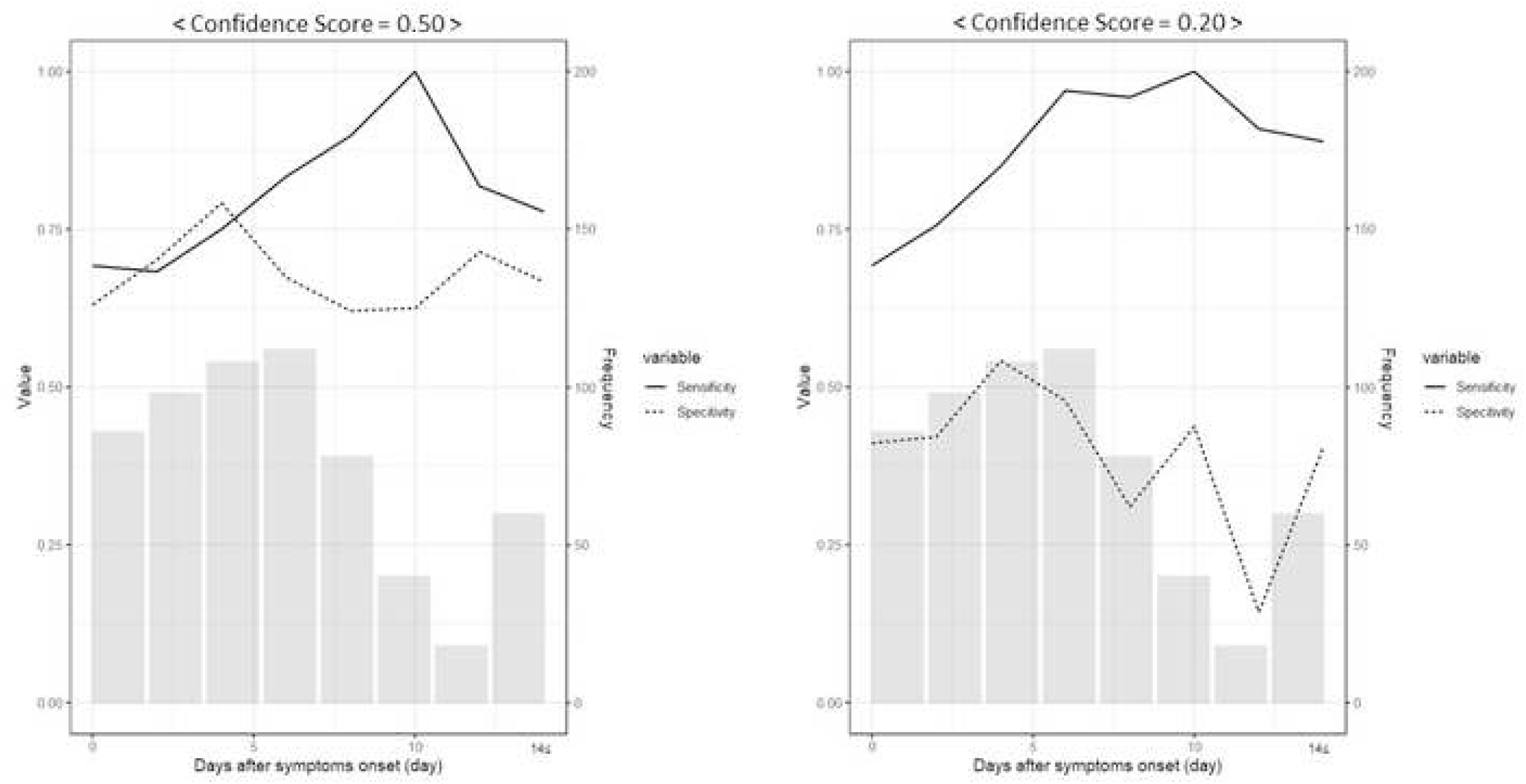
Differential performance of Ali-M3 for coronavirus disease in asymptomatic patients and patients using oxygen support. (A) A plot of test sensitivity (y-coordinate) versus its false-positive rate (x-coordinate) obtained at each cutoff level confidence score in asymptomatic patients. The area under the receiver operating characteristic (ROC) curve is 0.623 and the Youden index is 0.25. (B) A plot of test sensitivity, specificity, positive predictive value (PV+), and negative predictive value (PV-) in y-coordinate versus confidence score obtained from Ali-M3 in x coordinate among asymptomatic patients. The PV+ is dark gray and PV-is light gray. The maximum PV+ is 43.0% and maximum PV-is 57.0%. (C) A plot of test sensitivity (y-coordinate) versus its false-positive rate (x-coordinate) obtained at each cutoff level confidence score in patients using oxygen support. The area under the ROC curve is 0.623 and the Youden index is 0.25. (D) A plot of test sensitivity, specificity, PV+, and PV-in y-coordinate versus confidence score obtained from Ali-M3 in x-coordinate in patients using oxygen support. The PV+ is dark gray and the PV-is light gray. The maximum PV+ is 43.5% and the maximum PV-is 56.5%.

##### Dataset focused on asymptomatic patients

There were 86 asymptomatic patients (RT-PCR positive: 37). Using these patients only, the AUC was 0.623. When the cut-off point was 0.5, the sensitivity and specificity were 51.4% and 59.2%, respectively. When the cut-off point was 0.2, the sensitivity and specificity were 44.9% and 73.0%, respectively.

##### Dataset focused on patients needing oxygen therapy

There were 223 patients who required oxygen support (RT-PCR positive: 97). When using these patients only, the AUC was 0.828. When the cut-off point was set at 0.5, the sensitivity and specificity were 88.7% and 57.9%, respectively. When the cut-off point was set at 0.2, the sensitivity and specificity were 97.9% and 34.9%, respectively.

#### 5. Effect of the thickness of the CT reconstruction slice of CT

There were 320 patients (RT-PCR positive: 121) with a reconstruction slice thickness of under 3 mm When considering these patients only, the AUC was 0.825. When the cut-off point was set at 0.5, the sensitivity and specificity were 82.6% and 69.7%, respectively. When the cut-off point was set at 0.2, the sensitivity and specificity were 94.2% and 51.5%, respectively. In patients with a reconstruction slice thickness over 3 mm, the AUC was 0.789 (Supplement Figure 1)

## Discussion

In this external validation study, our results indicated that Ali-M3 could be useful for early triage of suspected COVID-19 patients with symptoms at a lower cut-off. In particular, higher accuracy was observed in patients with higher severity and a few days since symptom onset, and with images with a thinner reconstructed CT slice thickness.

Currently, all patients with symptoms, such as fever, are triaged as COVID-19 patients. Thus, medical practitioners must use PPE for each patient.[24] Additionally, bed zoning is essential to avoid contamination of non-infected patients.[25] On the other hand, under-triage cause hospital infections through admission of infected patients to facilities. This should continue until a definitive diagnosis is established. Since Ali-M3 is available on the cloud, the physician can receive the results immediately by sending the digital imaging and communications in medicine images from the ordinal picture archiving and communication system. When applying triage, clinicians require sufficient accuracy in terms of sensitivity, but specificity is less important.[19] The high sensitivity obtained at a cut-off of 0.2 with the AI diagnosis is useful for exclude the diagnosis of COVID-19.

Ali-M3 also has the potential to support a diagnosis of COVID-19. The tools currently used for diagnosing COVID-19 infection are antibody, antigen, and RT-PCR tests. Both antigen and RT-PCR tests use tracheal secretions or saliva. An antigen test requires an antigen protein above a given detectable level, and is currently inferior to RT-PCR tests. As the same patient sample is used, the antigen test cannot support the RT-PCR test. The RT-PCR test is currently used as a gold standard, but the sensitivity changes depending on the number of days after the onset of symptoms.[19] Therefore, for an exclusion diagnosis, multiple tests staggered over time are needed, rather than a single negative RT-PCR test. Even when this test is performed as rapidly as possible, it still requires a few days to obtain multiple test results. On the other hand, Ali-M3 uses the configurational information of patients’ lungs, and can add different information than obtained from RT-PCR, thereby complementing the drawbacks of RT-PCR.

In this study, the diagnostic accuracy at the validation stage was lower than the accuracy at the development stage. A two-gate (case-control) design was used in the development of the AI system but in the present study for evaluating the ability of Ali-M3 to assess a COVID-19 diagnosis by chest CT image, we used a single-gate (cohort) design. Although many studies have used the two-gate design in evaluation of AI for the diagnosis of COVID-19,[26] the two-gate design is generally prone to overestimation of diagnostic test results.[27] Thus, blindly using the results based on a two-gate design in a clinical situation can be inappropriate. Moreover, other factors should be considered. With the use of a two-gate design, the fact that RT-PCR is an imperfect reference standard is typically ignored. Furthermore, performing culture and tests to ascertain the true sensitivity of this test is difficult. In the present study, we simulated the diagnostic ability of Ali-M3 with consideration that the sensitivity of the reference standard was imperfect, which leads to underestimation of the specificity and AUC of Ali-M3, without distortion of the sensitivity. Furthermore, the outcomes while developing Ali-M3 and while examining its adequacy were different. Taking into account the patient flow in China, the outcomes at the development stage were set as positive cases with RT-PCR negative results and positive CT image findings.[28] This had a small effect on the sensitivity, but a large effect on the specificity. For example, if in the development stage, 33.9% of the positive patients had negative RT-PCR results and positive CT image findings,[28] then the performance that showed a sensitivity of 98.5% and specificity of 99.2% in the developing Ali-M3,[16] changes from 97.7% to 100% for sensitivity and from 80.8% to 81.6% for specificity when positive RT-PCR is the only reference used. Upgrading to a diagnostic AI that targets only RT-PCR-positive cases at the development stage is desirable.

This study had some limitations. First, the differentiation performance of Ali-M3 was poor in asymptomatic patients; thus, Ali-M3 should not be used to screen asymptomatic patients. While an alternative to the RT-PCR test for COVID-19 is expected in terms of screening for nosocomial infections and screening on admission for patients with other diseases, Ali-M3 is not recommended for this purpose. Second, we could not differentiate COVID-19 from other viral pneumonias. Compared to the past five seasons, the number of Japanese people infected with influenza during this season was markedly low.[29] In fact, only a few cases in our cohort were diagnosed with other viral pneumonias. Third, it could not reflect the difference in imaging features caused by different COVID-19 types. In addition to type A COVID-19 that was initially prevalent in Asia, type B and type C were prevalent in Europe and in the United States. These different types were not determined in the PCR test, and thus we could not evaluate these differences.

In conclusion, we conducted a retrospective cohort study for external validation of Ali-M3. Our results indicated that AI-based CT diagnosis could be useful for a diagnosis of exclusion of COVID-19 in symptomatic patients, particularly those requiring oxygen and with only a few days since symptom onset. Using Ali-M3 support can reduce PPE consumption and prevent hospital infections through the admission of covertly infected patients. Moreover, Ali-M3 also has the potential to support the diagnosis of RT-PCR for suspected COVID-19 patients. However, as Ali-M3 had some limitations in terms of development, further studies and learning are warranted for updating the system.

## Supporting information

STARD list

## Data Availability

The data that support the findings of this study are available from the corresponding author, Tatsuyoshi Ikenoue, upon permission of IRB at Hyogo Prefectural Amagasaki General Medical Center and reasonable request.

## Compliance with ethical standards

### Guarantor

The scientific guarantor of this publication is Tatsuyoshi Ikenoue.

### Funding

The authors state that this work has not received any funding.

### Statistics and biometry

No complex statistical methods were necessary for this paper.

### Informed consent

Written informed consent was waived by the Institutional Review Board.

### Ethical approval

Institutional Review Board approval was obtained.

### Study subjects or cohorts overlap

Any study subjects or cohorts have not been previously reported.

### Methodology

- retrospective
- diagnostic or prognostic study
- multicentre study

## Acknowledgments

We thank M3 Inc., and Clinical Porter for the support with providing free Ali-M3 and data storage, although they did not participate in the preparation protocol and manuscript. To want to access Ali-M3, reader can contact M3. We also thank Ms. Kyoko Wasai, who assisted retrieving data.

## Abbreviations

AI: artificial intelligence
COVID-19: coronavirus disease 2019
CT: computed tomography

