## Supplement for "Accuracy of deep learning based computed tomography diagnostic system of COVID-19: a consecutive sampling external validation cohort study"

**Supplemental material**

Methods regarding simulation of sensitivity, specificity, and AUC considering imperfect reference of RT-PCR

First, we calculated the sensitivity of CT-AI for true COVID-19(*Se_CT_*) and the specificity of CT-AI for true COVID-19 (*Sp_CT_*). We set *Σε_πχρ_* for the sensitivity of RT-PCR and set *1* for the specificity of RT-PCR. Supposed prevalence as
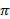

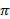
, true COVID-19 patients are $N\times\pi$ and true non-COVID-19 patients are
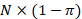
$N\times(1-\pi)$. We derived the 2*2 table as follows:

|  | | RT-PCR test | |
| --- | --- | --- | --- |
|  |  | Positive | Negative |
| CT-AI test | Positive | A | C |
|  | Negative | B | D |

*A = N*π*(Se_pcr_*Se_CT_) + N*(1-π)*((1-Sp_pcr_)*(1-Sp_CT_))*

*B = N*π*(Se_pcr_*(1-Se_CT_)) + N*(1-π)*((1-Sp_pcr_)*Sp_CT_)*

*C = N*π*((1-Se_pcr_)*Se_CT_) + N*(1-π)*(Sp_pcr_*(1-Sp_CT_))*

*D = N*π*((1-Se_pcr_)*(1-Se_CT_)) + N*(1-π)*(Sp_pcr_*Sp_CT_)*

When we substitute 1 for *Sp_pcr_*, the following results are obtained:

*A = N*π*(Se_pcr_*Se_CT_)*

*B = N*π*(Se_pcr_*(1-Se_CT_))*

*C = N*π*((1-Se_pcr_)*Se_CT_) + N*(1-π)*(1-Sp_CT_)*

*D = N*π*((1-Se_pcr_)*(1-Se_CT_)) + N*(1-π)*Sp_CT_*

Suppose
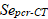
$\text{ }\text{Se}\text{pcr}\text{-CT }$ is the sensitivity of CT-AI when RT-PCR is established as the reference, the following results are obtained:

$$\text{Se}\text{pcr}\text{-CT }$$

$$\text{= }\frac{\text{A}}{\text{A+B}}\text{= }\frac{N\times\pi\times{(Se}_{pcr}\times{Se}_{CT})}{N\times\pi\times{(Se}_{pcr}\times{Se}_{CT})+N\times\pi\times{(Se}_{pcr}\times{(1-Se}_{CT}))}$$

=$\frac{{Se}_{pcr}\times{Se}_{CT}}{{Se}_{pcr}\times{Se}_{CT}+{Se}_{pcr}{-{Se}_{pcr}\times Se}_{CT}}= \frac{{Se}_{pcr}\times{Se}_{CT}}{{Se}_{pcr}}= {Se}_{CT}$

Hence, the sensitivity of CT-AI when RT-PCR is established as the reference is the true sensitivity of CT-AI for COVID-19 even if the sensitivity of RT-PCR is variable.

Supposed
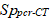
 $\text{S}\text{p}\text{pcr}\text{-CT }$ as the specificity of CT-AI when RT-PCR is set as reference, the following results are obtained:

$${Sp}_{pcr-CT}= \frac{D}{C+D}$$

$$= \frac{N\times\pi\left( \left( 1-{Se}_{pcr} \right)\times\left( 1-{Se}_{CT} \right) \right)+N\times(1-\pi)\times{Sp}_{CT}}{N\times\pi\left( \left( 1-{Se}_{pcr} \right)\times{Se}_{CT} \right)+N\times\left( 1-\pi\right)\times\left( 1-{Sp}_{CT} \right)+N\times\pi\left( \left( 1-{Se}_{pcr} \right)\times\left( 1-{Se}_{CT} \right) \right)+N\times(1-\pi)\times{Sp}_{CT}}$$

$$=\frac{\pi\left( \left( 1-{Se}_{pcr} \right)\times\left( 1-{Se}_{CT} \right) \right)+(1-\pi)\times{Sp}_{CT}}{1-\pi\times{Se}_{pcr}}$$

$$\therefore{Sp}_{CT}=\frac{{Sp}_{pcr-CT}\times\left( 1-\pi\times{Se}_{pcr} \right)-\pi\times\left( 1-{Se}_{pcr} \right)\times\left( 1-{Se}_{CT} \right)}{1-\pi}$$

Now, because the specificity of RT-PCR is *1,* we can derive another 2*2 table as follows:

|  | | True Disease | |  |
| --- | --- | --- | --- | --- |
|  |  | COVID-19 | Non-COVID-19 |  |
| RT-PCR test | Positive | X | 0 | X = N_PCR(+)_ |
|  | Negative | Y | Z | Y+Z = N_PCR(-)_ |
|  | | X+Y | Z | X+Y+Z = N |

Supposed $\text{Se}\text{pcr}$
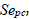
as the sensitivity of CT-AI when RT-PCR is set as reference, we derive the following:

$${Se}_{PCR}=\frac{X}{X+Y}$$

$$X+Y=\frac{N_{PCR(+)}}{{Se}_{PCR}}$$

$$\pi=\frac{X+Y}{X+Y+Z}=\frac{N_{PCR(+)}}{{Se}_{PCR}}\times\frac{1}{N}$$

$$\therefore{Sp}_{CT}=\frac{{Sp}_{pcr-CT}\times\left( 1-\pi\times{Se}_{pcr} \right)-\pi\times\left( 1-{Se}_{pcr} \right)\times\left( 1-{Se}_{CT} \right)}{1-\pi}$$

$$=\frac{\frac{N_{PCR(-)}}{N}\times{Sp}_{pcr-CT}-\frac{N_{PCR(+)}}{N}\times\frac{1}{{Se}_{pcr}}\times\left( 1-{Se}_{pcr} \right)\times\left( 1-{Se}_{pcr-CT} \right)}{\frac{N\times{Se}_{pcr}-N_{PCR(+)}}{N\times{Se}_{pcr}}}$$

$$=\frac{N_{PCR(-)}\times{Sp}_{pcr-CT}\times{Se}_{pcr}-N_{PCR(+)}\times\left( 1-{Se}_{pcr} \right)\times\left( 1-{Se}_{pcr-CT} \right)}{N\times{Se}_{pcr}-N_{PCR(+)}}$$

We simulated the ROC curve of CT-AI using this information and R. The maximum ${Se}_{pcr}$ is 1. We want to know the minimum ${Se}_{pcr}$. The lower ${Se}_{pcr}$, the higher the highest true specificity of CT-AI. However, the highest true specificity of CT-AI must be under 1. We used this information for R code. The R code is given below.

```
ROC0 <- function(    disease,
                        normal,
                        lowest=NULL,
                        width=NULL)
{
        my.hist <- function(x, brks)
        {
                k <- length(brks)
                freq <- numeric(k)
                for (i in 1:(k-1)) {
                        freq[i] <- sum(brks[i] <= x & x < brks[i+1])
                }
                freq[k] <- sum(x >= brks[k])
                freq
        }
        x <- c(disease, normal)
        min.x <- min(x)
        max.x <- max(x)
        if (is.null(lowest) || is.null(width)) {
                temp<- pretty(c(disease, normal), n=min(length(disease)+length(normal), 50))
                lowest <- temp[1]
                width <- diff(temp)[1]
        }
        brks <- seq(lowest, max.x+width, by=width)
        ROC(brks, my.hist(disease, brks), my.hist(normal, brks))
}

yoden_fun <- function(d)
{
sum = 0
best_sens  = 0
best_spec  = 0
best_point = 0

for (i in 1:nrow(d)){
    x = d[i,1]
    sens = d[i,2]
    spec = d[i,3]
    if((sens+spec)>sum){
        sum = sens+spec
        best_sens  = sens
        best_spec  = spec
        best_point = x
    }
}
return(list(best_point, best_sens, best_spec))
}

# 1.1  Crude
ROC <- function(     x,
                        disease,
                        normal)
{
        k <- length(x)
        stopifnot(k == length(disease) && k == length(normal))
        Sensitivity <- c(rev(cumsum(rev(disease)))/sum(disease), 0)
        False.Positive.Rate <- c(rev(cumsum(rev(normal)))/sum(normal), 0)
        Sp_pcrct <- 1-False.Positive.Rate

        plot(False.Positive.Rate, Sensitivity, type="b")
        abline(h=c(0, 1), v=c(0, 1))
        c.index <- sum(sapply(1:k, function(i)
(False.Positive.Rate[i]-False.Positive.Rate[i+1])*　(Sensitivity[i+1]+Sensitivity[i])/2))
# area under ROC curve
        result <- cbind(x, disease, normal, Sensitivity[-k-1], Sp_pcrct[-k-1], False.Positive.Rate[-k-1])
        rownames(result) <- as.character(1:k)
        colnames(result) <- c("Value", "Disease", "Normal",
"Sensitivity", "Specificity", "F.P. rate")

        d <- cbind(x, Sensitivity[-k-1], Sp_pcrct[-k-1])
        d <- yoden_fun(d)
        return(list(result=result, c.index=c.index, d=d))

}

disease.x <- dat %>%
  dplyr::filter(PCR == 1)
normal.x <- dat %>%
  dplyr::filter(PCR == 0)

ROC0(disease.x$Alibaba.Confidence, normal.x$Alibaba.Confidence)

# 1.2  Considering PCR Sensitivity
ROC <- function(     x,
                        disease,
                        normal)
{
        k <- length(x)
        stopifnot(k == length(disease) && k == length(normal))
        Sensitivity <- c(rev(cumsum(rev(disease)))/sum(disease), 0)
        False.Positive.Rate <- c(rev(cumsum(rev(normal)))/sum(normal), 0)

        total <- N
        pos   <- NPCR_ps
        nega  <- NPCR_ng

        for (i in 1:500){
        Sens_pcr <- 1 - 0.001 * i
        Sp_pcrct <- 1-False.Positive.Rate
        Specificity <- (nega*Sp_pcrct*Sens_pcr -pos*(1-Sens_pcr)*
(1-Sensitivity))/(total*Sens_pcr-pos)
        print(i)

        s = 0
        for (j in 1:50){
            if (Specificity[j] > s) {s <- Specificity[j]}
            }
```

if (s >= 1) [30]

```
        }
        Sens_pcr <- Sens_pcr + 0.001
        Sp_pcrct <- 1-False.Positive.Rate
        Specificity <- (nega*Sp_pcrct*Sens_pcr-pos*(1-Sens_pcr)*(1-Sensitivity))
/(total*Sens_pcr-pos)
        False.Positive.Rate <- 1-Specificity

        plot(False.Positive.Rate, Sensitivity, type="b")
        abline(h=c(0, 1), v=c(0, 1))
        c.index <- sum(sapply(1:k, function(i)
(False.Positive.Rate[i]-False.Positive.Rate[i+1])*　(Sensitivity[i+1]+Sensitivity[i])/2))
# area under ROC curve
        result <- cbind(x, disease, normal, Sensitivity[-k-1],
Specificity[-k-1], False.Positive.Rate[-k-1])
        rownames(result) <- as.character(1:k)
        colnames(result) <- c("Value", "Disease", "Normal", "Sensitivity",
"Specificity", "F.P. rate")

        d <- cbind(x, Sensitivity[-k-1], Specificity[-k-1])
        d <- yoden_fun(d)
        return(list(result=result, c.index=c.index, d=d))
}
disease.x <- dat %>%
  dplyr::filter(PCR == 1)
normal.x <- dat %>%
  dplyr::filter(PCR == 0)

ROC0(disease.x$Alibaba.Confidence, normal.x$Alibaba.Confidence)
```

Supplemental Table 1.

| **Section/Topic** | **Item** | **Checklist Item** | **Page** |
| --- | --- | --- | --- |
| **Title and abstract** | | | |
| Title | 1 | Identify the study as developing and/or validating a multivariable prediction model, the target population, and the outcome to be predicted. | Title page |
| Abstract | 2 | Provide a summary of objectives, study design, setting, participants, sample size, predictors, outcome, statistical analysis, results, and conclusions. | 1 |
| **Introduction** | | | |
| Background and objectives | 3a | Explain the medical context (including whether diagnostic or prognostic) and rationale for developing or validating the multivariable prediction model, including references to existing models. | 3 |
|  | 3b | Specify the objectives, including whether the study describes the development or validation of the model or both. | 3 |
| **Methods** | | | |
| Source of data | 4a | Describe the study design or source of data (e.g., randomized trial, cohort, or registry data), separately for the development and validation data sets, if applicable. | 4 |
|  | 4b | Specify the key study dates, including start of accrual; end of accrual; and, if applicable, end of follow-up. | 4 |
| Participants | 5a | Specify key elements of the study setting (e.g., primary care, secondary care, general population) including number and location of centers. | 4 |
|  | 5b | Describe eligibility criteria for participants. | 4 |
|  | 5c | Give details of treatments received, if relevant. | not applicable |
| Outcome | 6a | Clearly define the outcome that is predicted by the prediction model, including how and when assessed. | 5 |
|  | 6b | Report any actions to blind assessment of the outcome to be predicted. | 5 |
| Predictors | 7a | Clearly define all predictors used in developing or validating the multivariable prediction model, including how and when they were measured. | 5 |
|  | 7b | Report any actions to blind assessment of predictors for the outcome and other predictors. | 5 |
| Sample size | 8 | Explain how the study size was arrived at. | 4 |
| Missing data | 9 | Describe how missing data were handled (e.g., complete-case analysis, single imputation, multiple imputation) with details of any imputation method. | 5 |
| Statistical analysis methods | 10c | For validation, describe how the predictions were calculated. | 5 |
|  | 10d | Specify all measures used to assess model performance and, if relevant, to compare multiple models. | 5 |
|  | 10e | Describe any model updating (e.g., recalibration) arising from the validation, if done. | not applicable |
| Risk groups | 11 | Provide details on how risk groups were created, if done. | not applicable |
| Development vs. validation | 12 | For validation, identify any differences from the development data in setting, eligibility criteria, outcome, and predictors. | 4,5 |
| **Results** | | | |
| Participants | 13a | Describe the flow of participants through the study, including the number of participants with and without the outcome and, if applicable, a summary of the follow-up time. A diagram may be helpful. | 7  Figure 1 |
|  | 13b | Describe the characteristics of the participants (basic demographics, clinical features, available predictors), including the number of participants with missing data for predictors and outcome. | 7 Table 1 |
|  | 13c | For validation, show a comparison with the development data of the distribution of important variables (demographics, predictors and outcome). | Supplement Table 4 |
| Model performance | 16 | Report performance measures (with CIs) for the prediction model. | 7 |
| Model updating | 17 | If done, report the results from any model updating (i.e., model specification, model performance). | not applicable |
| **Discussion** | | | |
| Limitations | 18 | Discuss any limitations of the study (such as nonrepresentative sample, few events per predictor, missing data). | 11 |
| Interpretation | 19a | For validation, discuss the results with reference to performance in the development data, and any other validation data. | 10 |
|  | 19b | Give an overall interpretation of the results, considering objectives, limitations, results from similar studies, and other relevant evidence. | 10 |
| Implications | 20 | Discuss the potential clinical use of the model and implications for future research. | 9 |
| **Other information** | | | |
| Supplementary information | 21 | Provide information about the availability of supplementary resources, such as study protocol, Web calculator, and data sets. | Acknowledgment |
| Funding | 22 | Give the source of funding and the role of the funders for the present study. | Acknowledgment |

Supplemental Table 2. Inclusion criteria.

Patients who met the following criteria even for one item were considered symptomatic and were enrolled in the study.

| **Symptom** | **Level** |
| --- | --- |
| Fever | ≥ 37.0 C |
| Systolic blood pressure | ≤ 90 mmHg |
| Heart rate | ≥ 120 bpm |
| Respiratory rate | ≥ 25 /min |
| SpO2 | ≤ 92% |
| Use of catecholamine | Yes |
| Use of oxygen support | Yes |

Spo2, oxygen saturation; bpm, beats per minute

Supplemental Table 3. Computed tomography system and protocol.

| Facility | C01 | C02 | C03 | C04 | C05 | C06 | C07 | C08 | | C09 | C10 | C11 |
| --- | --- | --- | --- | --- | --- | --- | --- | --- | --- | --- | --- | --- |
| System | Aquilion PRIME | Optima CT660 | Aquilion PRIME | Optima CT660 | Optima CT660 | Aquilion PRIME | Aquilion CX Edition | Aquilion ONE | Aquilion CXL | Aquilion PRIME | Aquilion CXL | Aquilion CX Edition |
| Vendor | Canon Medical Systems | GE | Canon Medical Systems | GE | GE | Canon Medical Systems | Canon Medical Systems | Canon Medical Systems | Canon Medical Systems | Canon Medical Systems | Canon Medical Systems | Canon Medical Systems |
| Tube voltage (kVp) | 120 | 120 | 120 | 120 | 120 | 120 | 120 | 120 | 120 | 120 | 120 | 120 |
| Automatic tube current Modulation (mAs) | Auto | 100-510 | 150-250 | 80-500 | 80-500 | 150-250 | 403-500 | 100-400 | 100-400 | 50-250 | 100-400 | 100-400 |
| Pitch |  |  |  |  |  |  |  |  |  |  |  |  |
| Standard | 111 | 55 | 65 | 55 | 55 | 65 |  | 65 | 53 | 65 | 53 |  |
| Factor | 0.813 | 0.984 | 0.813 | 0.984 | 0.984 | 0.813 | 1.172 | 0.813 | 0.828 | 0.813 | 0.828 | 1 |
| Matrix | 512 × 512 | 512 × 512 | 256 × 256 | 512 × 512 | 512 × 512 | 512 ×512 | 512 ×512 | 512 × 512 | 512 × 512 | 512 × 512 | 512 × 512 | 512 × 512 |
| Slice thickness | 0.5 × 80 | 0.625 × 64 | 0.5 × 80 | 0.625 × 64 | 0.625 × 64 | 0.5 × 80 | 0.5 × 64 | 0.5 × 80 | 0.5 × 64 | 0.5 × 80 | 1.0 × 64 | 5.0 × 64 |
| Field of view (mm) | 320 | 340 | 320 |  |  | 330 | 350 | 320 | 320 | 320-400 | 320 | 320 |
| Reconstruction interval (mm) | 5 | 0.625 | 2 | 1.25 | 1.25 | 3 | 5 | 5 | 5 | 5 | 5 | 5 |

Supplement Table 4. Population characteristics in development from datasheet. [16]

| Category | | Number (%) | |
| --- | --- | --- | --- |
| ***Diagnosis*** | COVID-19 | 3,722 | (42.9) |
|  | Pneumonia | 2,491 | (28.7) |
|  | Non-pneumonia | 2,454 | (28.3) |
| ***Sex*** | Male | 4,162 | (48.0) |
|  | Female | 3,826 | (44.1) |
| ***Age*** | ≤50 years | 3,137 | (36.1) |
|  | > 50 years | 4,851 | (55.9) |

COVID-19, coronavirus disease
