## Supplementary figures and images for "Accuracy of deep learning based computed tomography diagnostic system of COVID-19: a consecutive sampling external validation cohort study"

### Supplement Figure

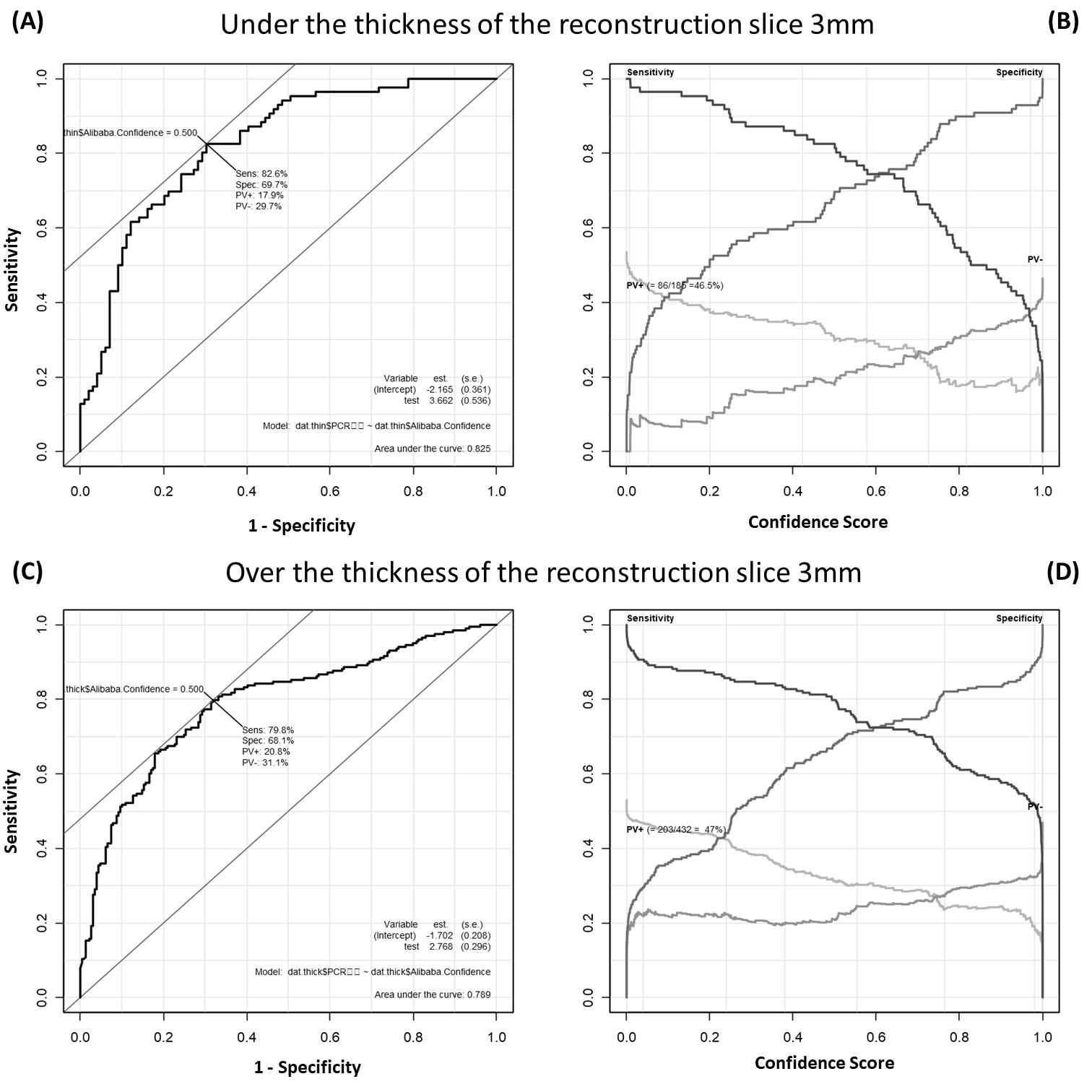
