## Supplement Figure's Legend for "Accuracy of deep learning based computed tomography diagnostic system of COVID-19: a consecutive sampling external validation cohort study"

Supplemental Figures

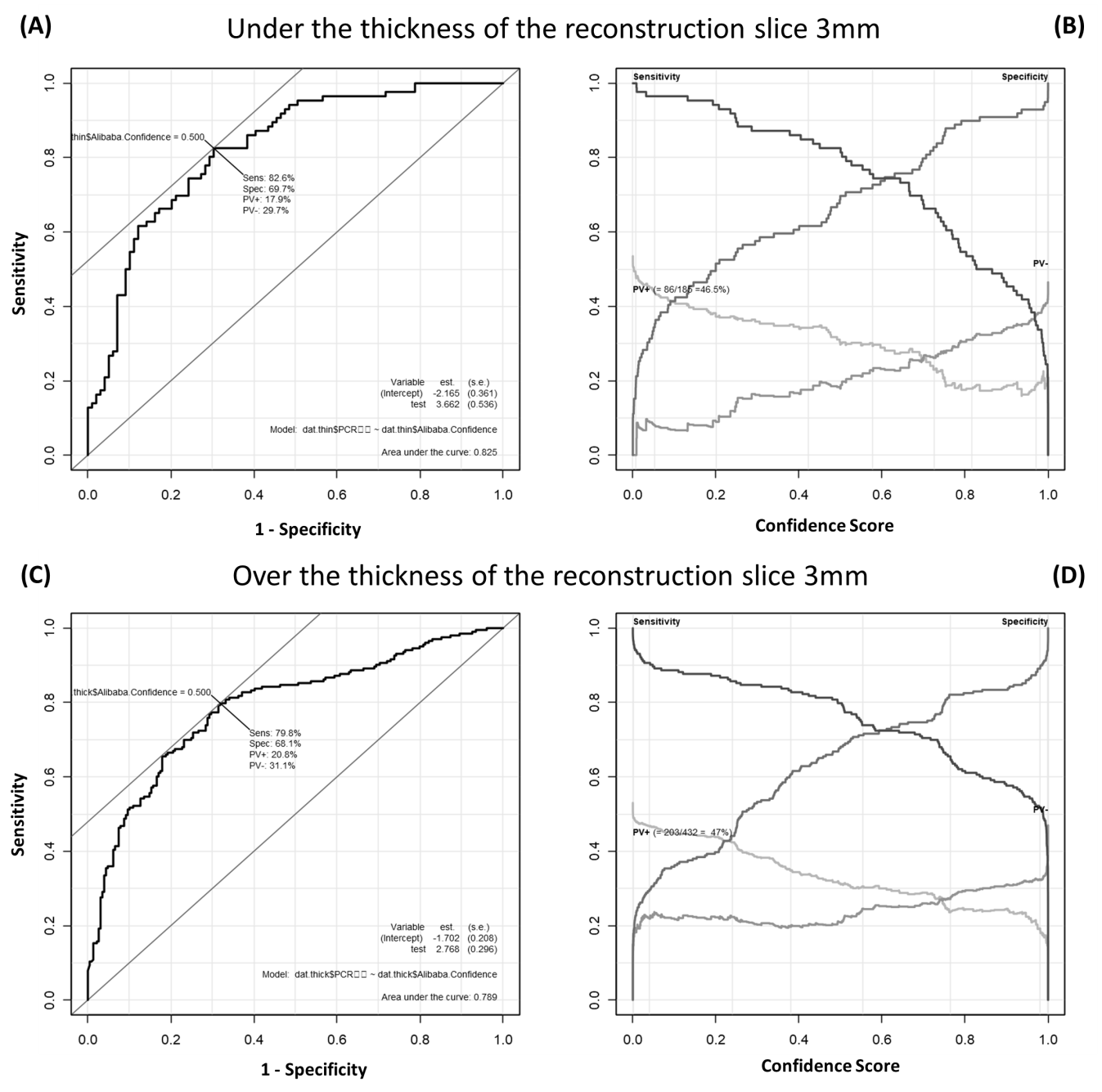

Supplement Figure 1. Differential performance of Ali-M3 for coronavirus disease in patients who were divided by the thickness of the reconstruction slice of computed tomography.

(A) A plot of test sensitivity (y coordinate) versus its false-positive rate (x coordinate) obtained at each cutoff level confidence score under the 3 mm thickness of the reconstruction slice. The area under the receiver operating characteristic (ROC) curve is 0.825 and the Youden index is 0.50. (B) A plot of test sensitivity, specificity, positive predictive value (PV+), and negative predictive value (PV-) in y coordinate versus confidence score obtained from Ali-M3 in x coordinate under the 3 mm thickness of the reconstruction slice. The PV+ is dark gray and the PV- is light gray. The maximum PV+ is 46.5% and the maximum PV- is 53.5%. (C) A plot of test sensitivity (y coordinate) versus its false-positive rate (x coordinate) obtained at each cutoff level confidence score over the 3 mm thickness of the reconstruction slice. The area under the ROC curve is 0.789 and the Youden index is 0.50. (D) A plot of test sensitivity, specificity, PV+, and PV- in y coordinate versus confidence score obtained from Ali-M3 in x coordinate over the 3 mm thickness of the reconstruction slice. The PV+ is dark gray and the PV- is light gray. The maximum PV+ is 47.0% and the maximum PV- is 53.0%.
